# Health Impact Assessment of PM_2.5_ attributable mortality from the September 2020 Washington State Wildfire Smoke Episode

**DOI:** 10.1101/2020.09.19.20197921

**Authors:** Yisi Liu, Elena Austin, Jianbang Xiang, Tim Gould, Tim Larson, Edmund Seto

## Abstract

Major wildfires that started in the summer of 2020 along the west coast of the U.S. have made PM_2.5_ concentrations in cities in this region rank among the highest in the world. Regions of Washington were impacted by active wildfires in the state, and by aged wood smoke transported from fires in Oregon and California. This study aims to assess the population health impact of increased PM_2.5_ concentrations attributable to the wildfire. Average daily PM_2.5_ concentrations for each county before and during the 2020 Washington wildfire episode were obtained from the Washington Department of Ecology. Utilizing previously established associations of short-term mortality for PM_2.5_, we estimated excess mortality for Washington attributable to the increased PM_2.5_ levels. On average, PM_2.5_ concentrations increased 91.7 μg/m^3^ during the wildfire episode. Each week of wildfire smoke exposures was estimated to result in 87.6 (95% CI: 70.9, 103.1) cases of increased all-cause mortality, 19.1 (95% CI: 10.0, 28.2) increased cardiovascular disease deaths, and 9.4 (95% CI: 5.1, 13.5) increased respiratory disease deaths. Because wildfire smoke episodes are likely to continue impacting the Pacific Northwest in future years, continued preparedness and mitigations to reduce exposures to wildfire smoke are necessary to avoid this excess health burden.

## 1. Introduction

A series of major wildfires has impacted air quality in western regions of the US in 2020, starting notably in Northern California on August 19, sparked by intense thunderstorms combined with a dry winter and sweltering summer. The northern California fires were followed by large fire complexes in Southern California, Oregon, and Washington. As of September 16, 22 large fires have been reported in California (2.3 million acres), 12 in Oregon (0.9 million acres), and 11 in Washington (0.7 million acres) ^1^. The intense smoke from these fires has resulted in major regional increases in concentrations of airborne particulates with a diameter of 2.5 μm or less (PM_2.5_). As winds shifted from fire-prone dry easterly flow to a northwesterly flow, smoke from the west coast fires that had traveled over the Pacific Ocean was transported to the Pacific Northwest. This has resulted in some of the highest air pollution levels ever observed in Washington.

Wildfire smoke consists of a mixture of air pollutants, including particulate matter, carbon monoxide, nitrogen oxides, acrolein, formaldehyde, benzene, benzo[a]pyrene, and dibenz[a,h]anthracene. The composition can vary from fire to fire, depending on the fuel (the type of vegetation, whether it burned through a town or structures), temperature and aging in the atmosphere ^2^. The particle size of the smoke tends to be small, approximately 0.4 to 0.7 μm in diameter. These particle sizes are harmful to human health because they are sufficiently small to be inhaled deep into the lungs ^2^. However, studies have not documented the increased PM_2.5_ concentrations, nor estimated the health impacts of this 2020 wildfire smoke episode in Washington.

The goal of this paper is to assess the potential human health impact of increased PM_2.5_ concentrations attributable to the wildfire smoke episode in 2020, which is the worst one on record for wildfires on the West Coast ^3^. As of the time of this analysis (September 30), major fires are ongoing along the western U.S. Therefore, the impact estimates are meant to help prepare for, and inform proactive measures to reduce smoke-induced health effects of this current smoke episode as wind patterns continue to carry smoke into other regions. The analysis utilizes evidence from previously published studies on short-term mortality associated with PM_2.5_, and concentration-response relationships. We estimate increases in all-cause, respiratory, and cardiovascular mortality for Washington, and discuss public health response activities carried out by air quality authorities.

## 2. Methods

### 2.1 PM_2.5_ Exposures

We obtained the daily PM_2.5_ concentration in each county from Washington’s Air Monitoring Network (https://enviwa.ecology.wa.gov/Report/Hr24PM25SummaryNew). Counties with regulatory air monitors were assigned the average of PM_2.5_ concentrations from available monitors in each county. Counties without regulatory monitors (six counties: Douglas County, Lincoln County, Pend Oreille County, San Juan County, Skamania County and Wahkiakum County) were assigned the average of PM_2.5_ concentrations of nearby counties. The PM_2.5_ levels before the wildfire were defined as the mean of daily PM_2.5_ concentrations from January 1 to September 6, 2020; PM_2.5_ levels during the wildfire period were defined as the mean of daily PM_2.5_ concentrations from September 7 to September 19, 2020.

### 2.2 Health Impact Assessment

We performed a health impact assessment for Washington to estimate the excess mortality attributable to increased PM_2.5_ during the wildfire smoke episode. The attributable fraction (AF) method was used to estimate the increased daily mortality following the equations below:

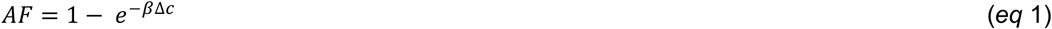

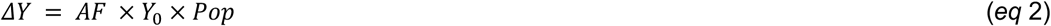

where AF is the attributable fraction of health events attributable to PM_2.5_ exposure, β is the cause-specific coefficient of the concentration-response functions (CRF) for PM_2.5_, *Δ*c is the change in PM_2.5_ due to the wildfire, *Δ*Y is the estimated health impact of PM_2.5_ attributable to PM_2.5_, Y_0_ is the baseline cause-specific mortality, and Pop is the exposed population.

The coefficient of CRF for short-term ambient air PM_2.5_ concentrations and mortality (including all-cause mortality, cardiovascular diseases mortality and respiratory diseases mortality) was adopted from a previous meta-analysis ^4^. The changes in PM_2.5_ before and during the wildfire was calculated by county as described above. The age-adjusted cause-specific mortality rate for the general population in Washington was obtained from the Washington State Department of Health ^5^. The resident population data for each county in Washington was the population estimates in 2019 from the U.S. Census Bureau ^6^.

After the above calculation by county, we summed the daily increased mortality for each county to calculate the increased daily deaths in Washington during the wildfire. Finally, we assessed the total increased deaths in Washington attributable to wildfire by multiplying the increase daily deaths by the total number of days of elevated exposure due to the wildfire episode.

### 2.3 Sensitivity Analysis

Nephelometers are used at different monitoring sites in the regulatory air quality monitoring network in Washington. Although they are normally calibrated to U.S. EPA federal reference method measurements, there is the potential that these calibrations may not be appropriate for the unusually high PM_2.5_ concentrations observed during the wildfire smoke episode. Therefore, we conducted an additional sensitivity analysis, computing health impacts with or without the PM_2.5_ concentrations reported from nephelometers. Since only 10 counties had daily PM_2.5_ concentrations after excluding data reported by nephelometers, the rest 29 counties were assigned the average of PM_2.5_ concentrations from all the included monitors in Washington. Following the same health impact assessment procedures, increased mortality attributable to wildfire in Washington was estimated for varying number of days of duration of wildfire smoke episode.

## 3. Results and Discussion

Figure A1 shows PM_2.5_ concentrations in selected counties before, during and after the 2020 wildfire smoke episode in Washington. PM_2.5_ levels started to climb since September 7, and were backed to normal around September 19-20. The average increment in PM_2.5_ concentration during the wildfire smoke episode was 91.7 μg/m^3^, with large variations (standard deviation, SD = 32.2 μg/m^3^) (Table 1). After excluding PM_2.5_ concentrations measured by nephelometers, the increase in PM_2.5_ levels reached 120.2 μg/m^3^ (SD: 54.9 μg/m^3^) (Table A1). Kittitas county located in central Washington was heavily impacted by smoke, where PM_2.5_ concentrations showed a 100-fold of increase over pre-wildfire levels (Table A2, A3).

**Table 1.** Summary of PM_2.5_ concentrations in Washington before and during the wildfires, 2020.

| PM<sub>2.5</sub> concentrations (µg/m<sup>3</sup>) | Mean | SD | Median | Range | Minimum | Maximum |
| --- | --- | --- | --- | --- | --- | --- |
| Before wildfire | 4.3 | 1.4 | 4.1 | 6.8 | 1.9 | 8.6 |
| During wildfire | 96.0 | 32.2 | 91.4 | 164.5 | 30.5 | 195.0 |
| Increases | 91.7 | 32.1 | 86.5 | 164.8 | 28.3 | 193.1 |

According to our health impact assessment, each week of wildfire smoke exposure at the measured ambient air concentrations would lead to 87.6 (95% CI: 70.9, 103.1) cases of increased all-cause mortality, 19.1 (95% CI: 10.0, 28.2) increased cardiovascular disease deaths, and 9.4 (95% CI: 5.1, 13.5) increased respiratory disease deaths. That is, 8.6% (95%CI: 7.0%, 10.2%) more all-cause deaths, 6.9% (95%CI: 3.6%, 10.2%) more cardiovascular diseases deaths, and 9.4% (95%CI: 5.1%, 13.5%) more respiratory diseases deaths occurred during this wildfire smoke episode compared to typical weeks. If the wildfire smoke episode lasts for a month (31 days), approximately 400 excess all-cause deaths would be attributable to the wildfire (Figure 1). The mortality burden of wildfire smoke exposure was the largest in King county, due to it having the largest population (Table A2 and Figure A2); whereas, Kittitas county showed the highest increased mortality rate (increased deaths per 100,000 persons) due to the largest increment of PM_2.5_ concentrations (Table A3 and Figure A2). The sensitivity analysis, excluding PM_2.5_ concentrations measured by nephelometers, resulted in slightly higher estimates, with each week of wildfire causing 102.8 (95% CI: 83.3, 120.8) more all-cause deaths, 22.5 (95% CI: 11.8, 33.0) increased cardiovascular disease deaths, and 11.0 (95% CI: 6.1, 15.7) excess respiratory disease deaths (Figure A3).

**Figure 1.**
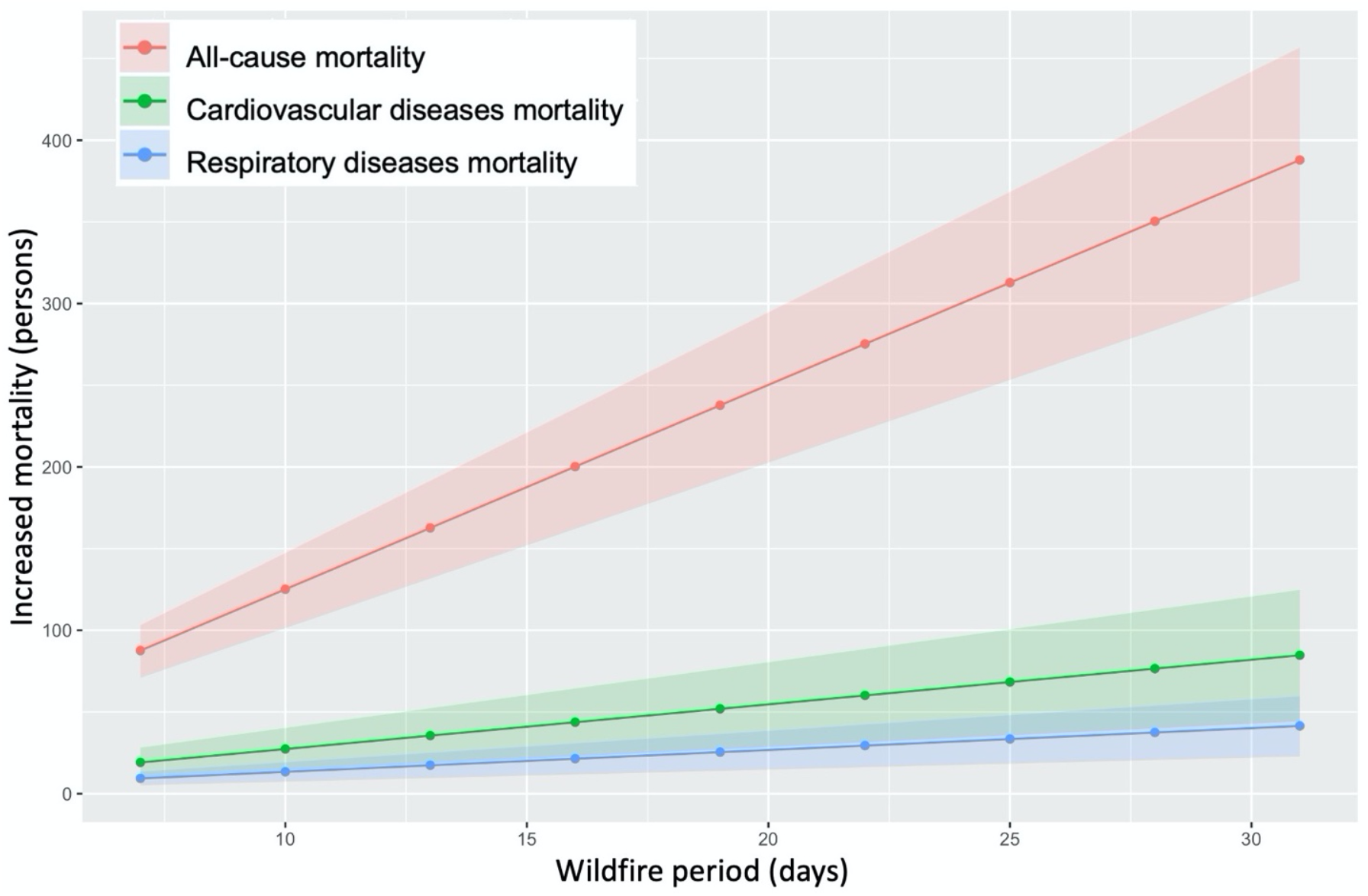
Increased mortality attributable to the wildfire in Washington.

Our findings indicate considerable excess mortality due to elevated PM_2.5_ concentrations that will increase with longer durations of the wildfire smoke episode in Washington. We estimate increases of 87.6 (95% CI: 70.9, 103.1) all-cause, 19.1 (95% CI: 10.0, 28.2) cardiovascular, and 9.4 (95% CI: 5.1, 13.5) respiratory deaths for each additional week of elevated wildfire smoke based on the average PM_2.5_ concentrations observed thus far. We obtained the daily PM_2.5_ concentration for each county from Washington’s Air Monitoring Network instead of EPA Air Quality System, because states flag data that may have been influenced by an exceptional event (e.g. wildfire, high winds) when reporting monitoring data to EPA. That is, PM_2.5_ concentrations obtained from EPA data repositories tend to be lower than that from state data repositories. Although our analysis focused on quantifying increased mortality, the findings are consistent with recent reports of increased hospital emergency room visits (increases of 3 people/day with respiratory issues at Harborview Medical Center, one of the larger hospitals in the Seattle area) and excess numbers of emergency medical service (EMS) calls in Snohomish County ^7^. While the news reporting has focused on respiratory issues, our findings indicate that with respect to mortality, the cardiovascular health effects may be greater than the respiratory effects.

Our findings are also consistent with previous studies of the health effects associated with wildfire smoke exposure. Systematic reviews provide consistent evidence for the positive associations between wildfire smoke exposures and all-cause mortality and respiratory health ^8-10^. A recent case-crossover study in Washington found the previous day’s exposure to wildfire smoke increased non-traumatic mortality by 2% (95% CI: 0.0, 5.0%), and the same-day exposure to wildfire smoke increased respiratory mortality by 9% (95% CI: 0.0, 18.0%) compared to non-wildfire smoke days ^11^. Although results are mixed for the cardiovascular health effects of wildfire smoke exposures, an increasing number of recent publications provide cumulative evidence for the association between wildfire smoke exposures and adverse cardiovascular outcomes. During the 2015 California wildfire, increased all-cause cardiovascular and cerebrovascular emergency department visits were reported on smoke days compared to days without smoke, especially on dense smoke days and among older adults (residents aged ≥65 years) ^12^. Similarly, another study found increased out-of-hospital cardiac arrest risk was associated with heavy smoke days (lag day 2 odds ratio: 1.7, 95% CI: 1.18, 2.13) in California from 2015 to 2017 ^13^. Additionally, a national study for more than 600 counties in the U.S. found cardiovascular hospitalizations increased 0.61% (95% CI: 0.09, 1.14) for each 10 μg/m^3^ increase in PM_2.5_ during smoke days ^14^.

The wildfire smoke episode in Washington ended on September 20th, 2020, which was estimated to result in approximately 163, 36 and 17 cases of increased all-cause, cardiovascular disease and respiratory disease deaths, respectively. The short-term excess mortality associated with this wildfire episode is large due to the relatively large increase in PM_2.5_ concentrations, not commonly observed in this part of the world. While aged wood smoke is still transporting to Washington from ongoing wildfires in California ^15^, and wildfire smoke episodes are likely to continue impacting the Pacific Northwest in future years, steps should be taken to reduce exposures to wildfire smoke to avoid this excess health burden.

Already, the state governor’s office has released a series of proclamations ^16^ aimed at addressing the impacts of this particular wildfire episode, including (1) declaring a state of emergency (20-86) that acknowledge the “threat to life and property… impacting the life and health of citizens… a public disaster demanding immediate action”, which activated the state’s Emergency Management Plan and directed resources to address the wildfire; (2) Providing financial assistance to individuals affected by the wildfires (20-72); (3) and maintaining critical firefighting supply lines and restoration of critical utilities (20-73 and 20-73.1). Additionally, several agencies (Department of Ecology, Department of Health, US Forest Service, regional Air Quality Agencies, County Health Departments, and Tribes) had collaborated in previous years to provide information to the public via a website ^17^, which includes material for this wildfire event, such as information on the health effects of wildfire smoke, a real-time map of regional air quality, guidance on both the US EPA air quality index (AQI) and the Washington state air quality advisory (WAQA) ^18^, and recommendations with respect to recognizing relevant health symptoms, acknowledgment of particularly susceptible subpopulations, reducing outdoor physical activity, staying indoors and implementing filtration strategies, mask use, and specific guides for schools, workers, parents of children, and information in Spanish. These activities serve as a good model for preparedness and response to address the health effects of wildfire smoke episodes.

While wildfire smoke episodes have impacted the Pacific Northwest region historically, this particular episode may be especially problematic due to the coincidence with the COVID-19 pandemic. Although there is still insufficient evidence to support this, exposure to elevated wildfire smoke may exacerbate the effects of SARS-CoV-2 infection. There is however clear evidence of increased acute lower respiratory infections with PM_2.5_ exposure, especially for children ^19-21^. Evidence is also available for the delayed effect of higher PM_2.5_ levels during the wildfire season on increased influenza in the following winter flu season ^22^. Given the current stay-at-home order to prevent the spread of SARS-CoV-2, fewer people may be outdoors and exposed to ambient PM_2.5_ than if there was not a pandemic. For instance, many children are not currently attending school in-person, which may result in less exposure to outdoor PM. However, essential workers, including many outdoor worker groups (e.g., firefighters, emergency personnel, agriculture and construction workers, delivery persons, etc.) may be groups that are more exposed to air pollution during this period, and thus might be at higher risk for health effects.

Some potential limitations of this analysis include first, the reliance on general PM_2.5_ mortality concentration-response functions rather than wildfire smoke-specific health effects. However, we have deemed this appropriate as they are based on evidence from a larger group of air pollution studies, and are within the range of effects reported for wildfire studies. Moreover, a previous study that assessed wildfire smoke impacts on hospitalizations in the U.S. did not find a difference between wildfire smoke PM_2.5_ vs non-wildfire smoke PM_2.5_-related increases in hospitalization ^14^; another study in Finland also showed that the associations between PM_2.5_ from wildfire and daily mortality were similar to the more general estimates of urban PM_2.5_ ^23^. Second, we did not employ highly spatially refined exposure modeling in this analysis, but we have considered county-specific variations. However, this scale is reasonable given the size of the smoke plume from long-range transport. Also, we have not yet assessed changes in state vital records, which we would anticipate to confirm the impact assessment estimates. However, this may be a useful future activity after the smoke episode has resolved. Finally, we have only assessed mortality impacts. This is probably just the “tip of the flame”, as there may be many more affected by PM_2.5_ related morbidity.

## Data Availability

The data that support the findings of this study are openly available in Washington Department of Ecology (https://enviwa.ecology.wa.gov/Report/Hr24PM25SummaryNew), and the U.S. Census Bureau (https://www.census.gov/data/datasets/time-series/demo/popest/2010s-counties-total.html).

## Acknowledgement

We wish to acknowledge the Washington Department of Ecology for its readily available PM2.5 concentration data from throughout its extensive statewide monitoring network.

## Appendix

**Table A1.** Summary of PM_2.5_ concentrations in Washington (excluding monitoring data from Nephelometers) before and during the wildfire episode, 2020.

| PM<sub>2.5</sub> concentrations ($\mu\text{g}/\text{m}^3$) | Mean | SD | Median | Range | Minimum | Maximum |
| --- | --- | --- | --- | --- | --- | --- |
| before wildfire | 5.4 | 2.5 | 5.4 | 10.1 | 0.4 | 10.5 |
| during wildfire | 125.6 | 54.0 | 118.2 | 186.0 | 49.3 | 235.3 |
| increases | 120.2 | 54.9 | 112.8 | 191.8 | 43.1 | 234.9 |

**Table A2.**
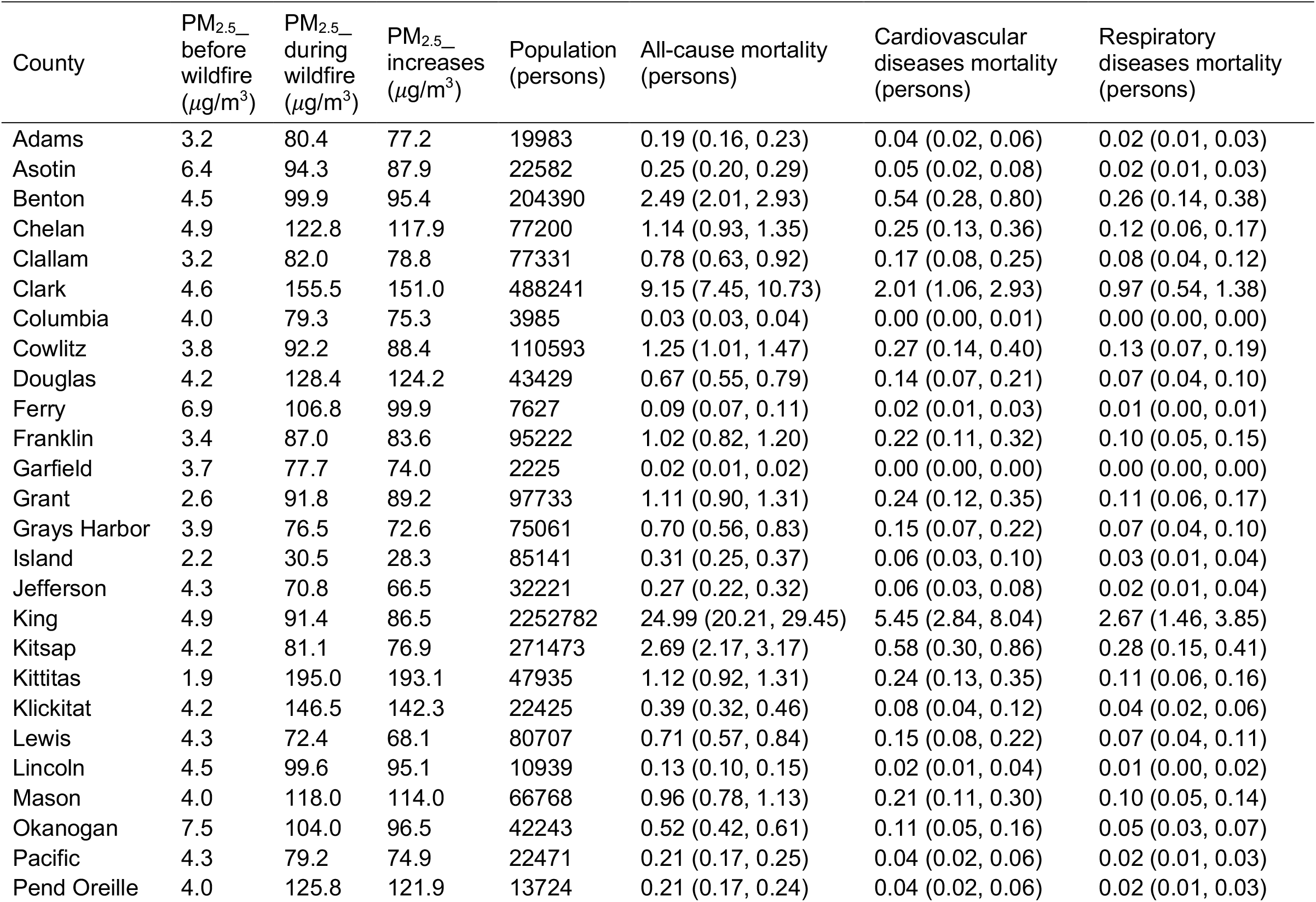
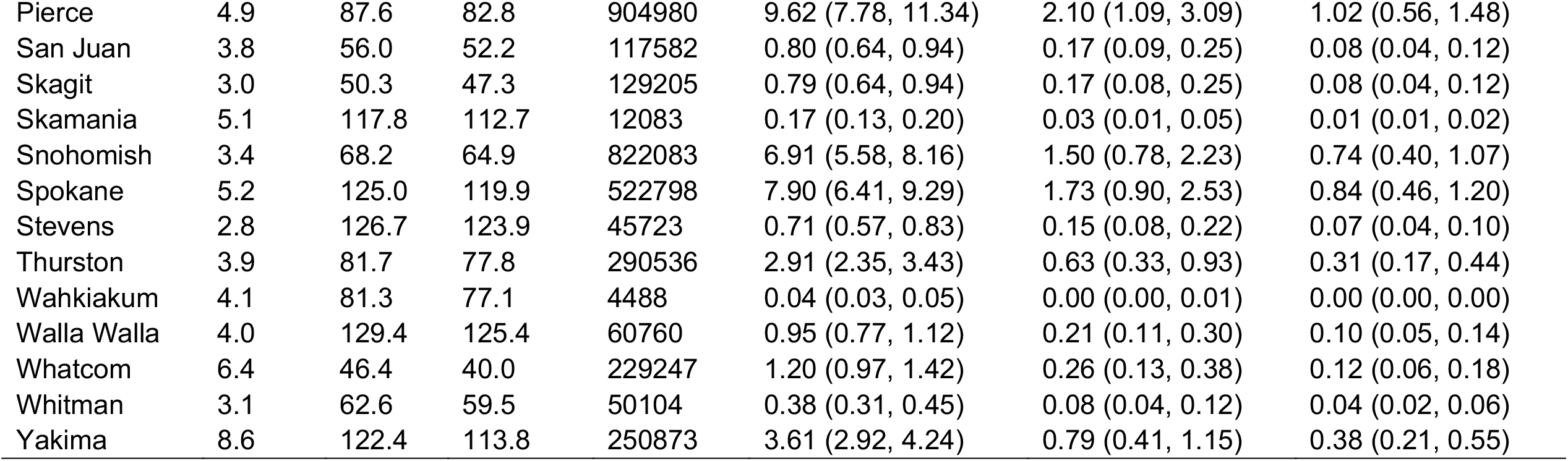
The average PM_2.5_ concentration changes in the wildfire episode and estimated mortality during each week of the wildfire episode for each county.

**Table A3.**
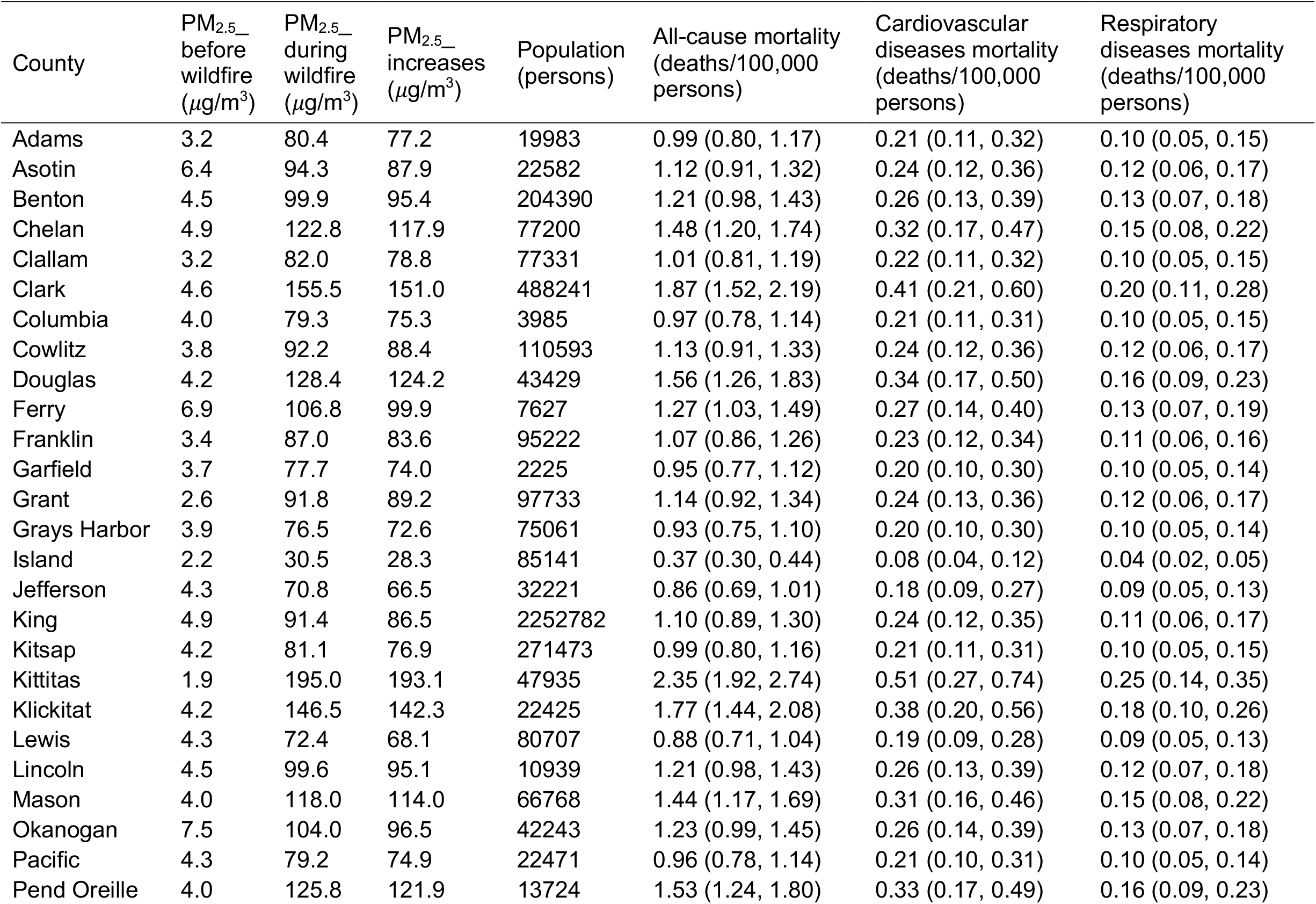
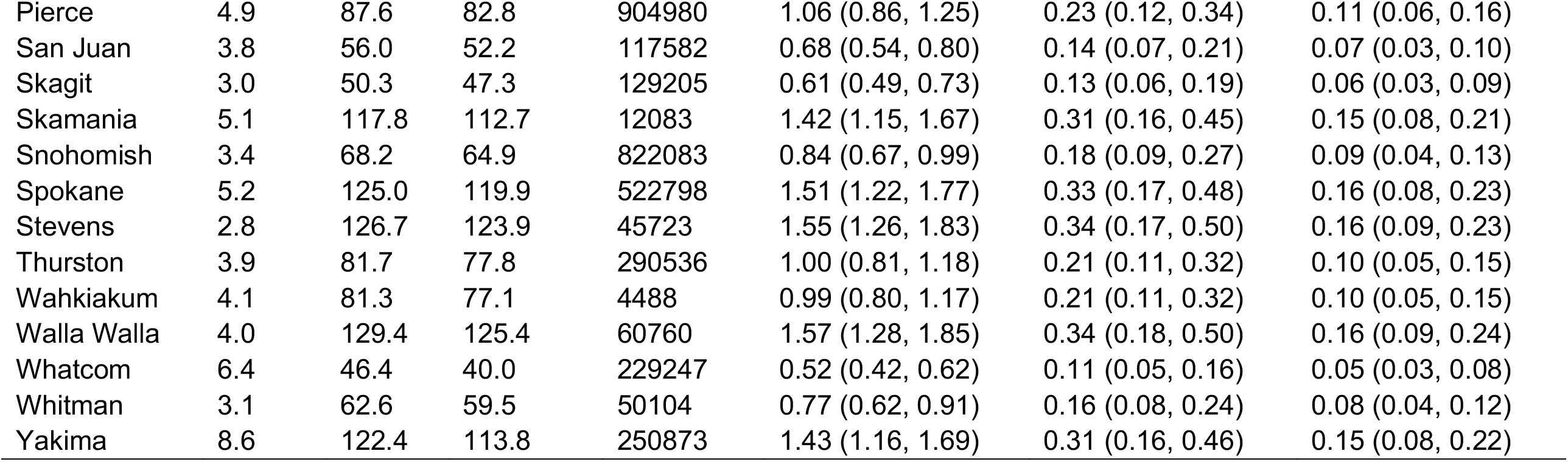
The average PM_2.5_ concentrations changes in the wildfire episode and estimated mortality per 100,000 persons during each week of the wildfire episode for each county.

**Figure A1.**
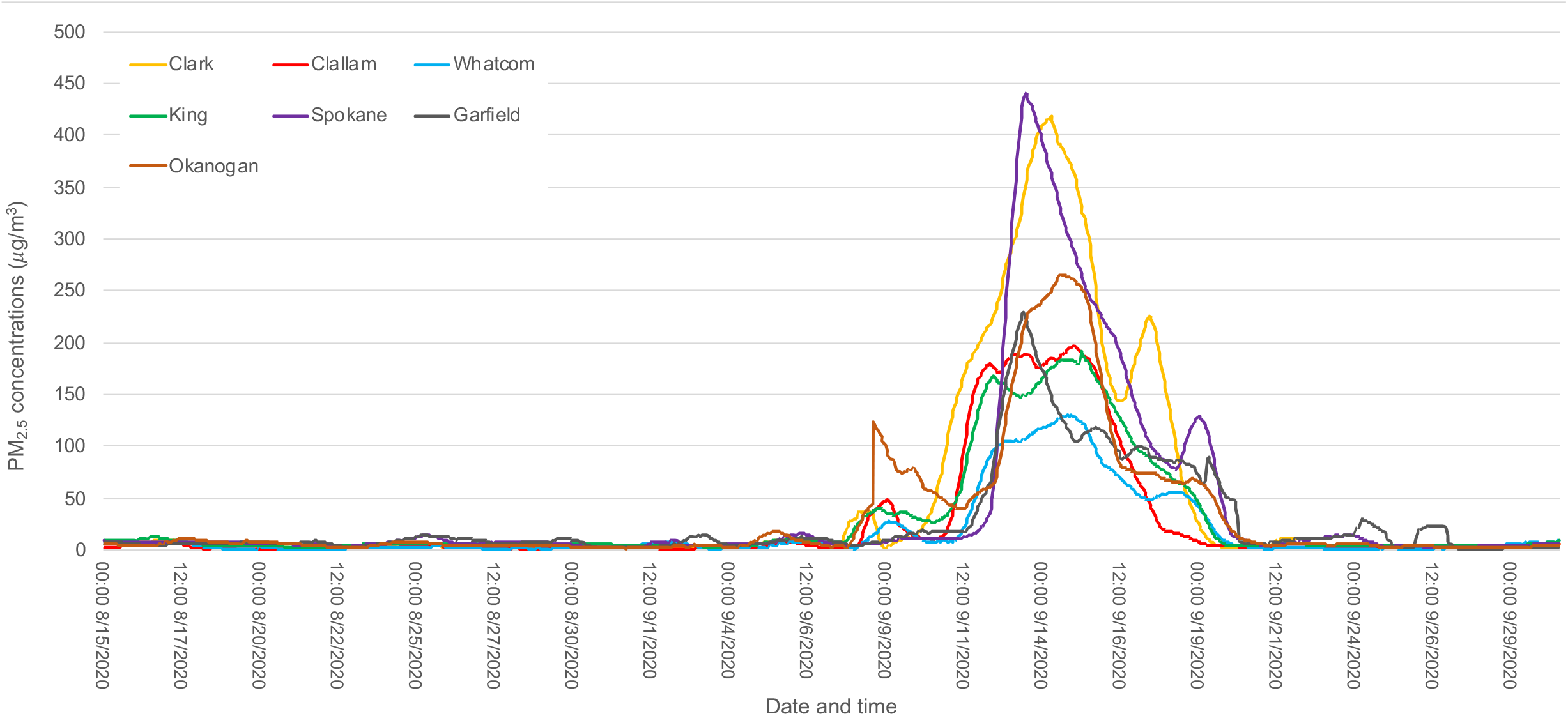
Time series plot of PM_2.5_ concentrations in selected counties before, during and after the 2020 wildfire smoke episode in Washington.

**Figure A2.**
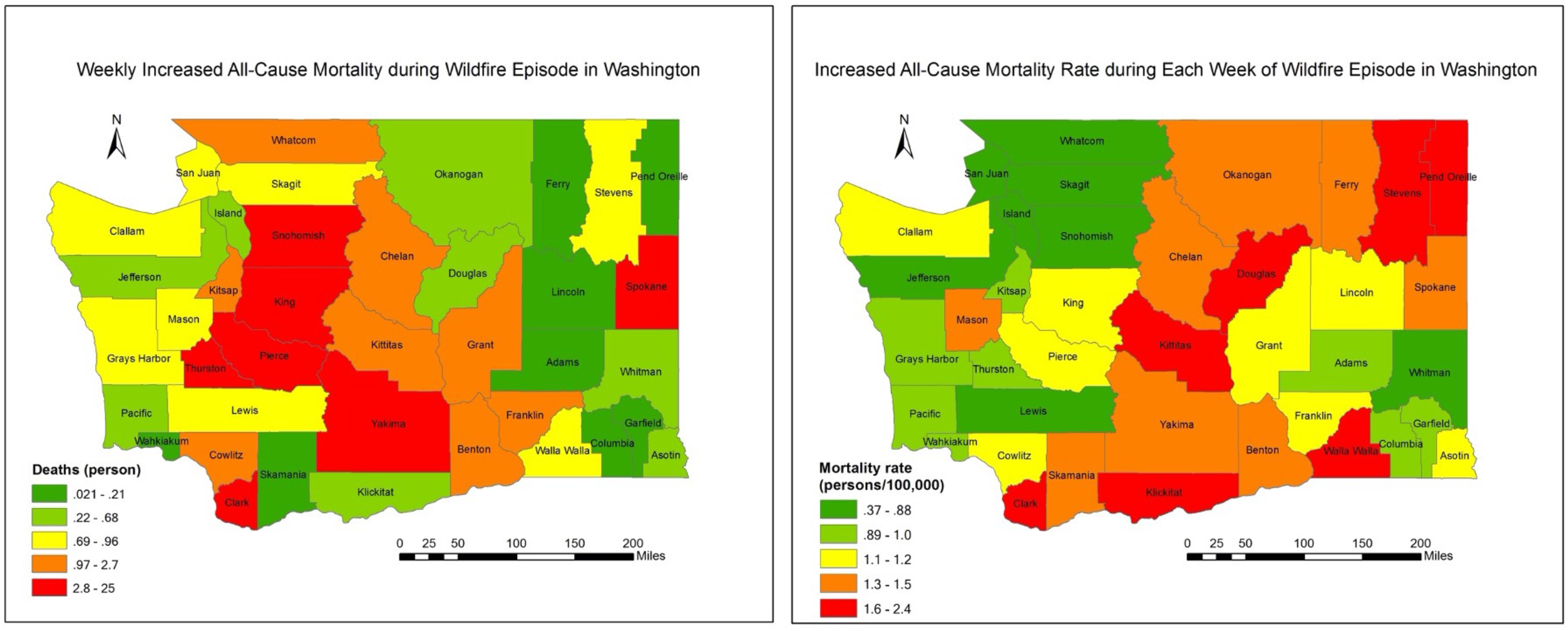
Mortality burden of each week of wildfire smoke exposure for each county in Washington.

**Figure A3.**
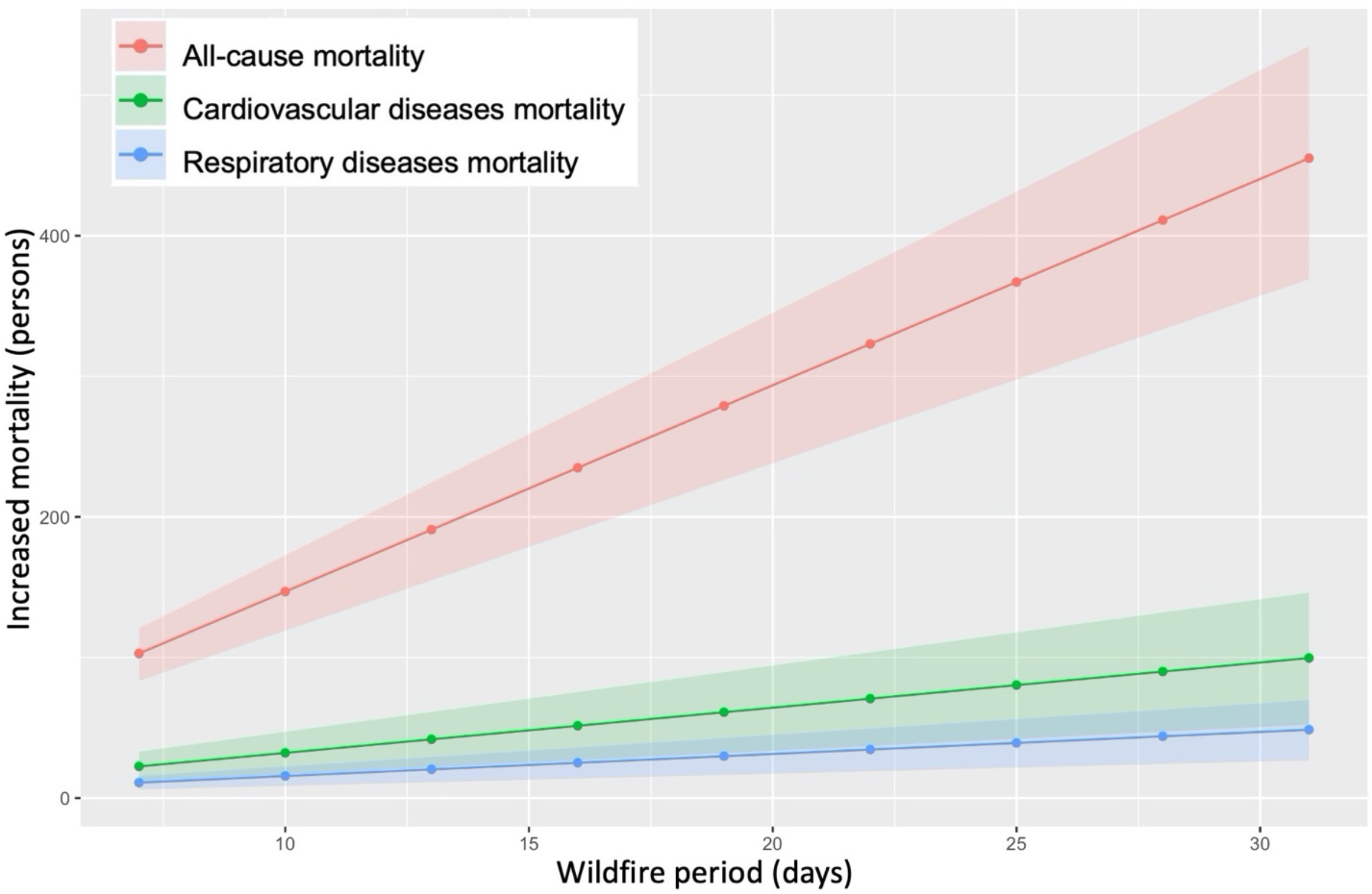
Increased mortality attributable to the wildfire in Washington (excluding monitoring data from Nephelometers).

